# Exploring the Relationship of Rush Hour Period and Fatal and Non-Fatal Crash Injuries in The U.S.: A Systematic Review and Meta-Analysis

**DOI:** 10.1101/2021.08.03.21261572

**Authors:** Oluwaseun John Adeyemi, Ahmed Arif, Rajib Paul

## Abstract

Road crashes are preventable causes of morbidity and mortality. In the U.S., substantial crashes occur during the rush hour period. The rush hour represents the period of the day during which the density of humans and vehicles in the road environment is highest. In the U.S., the rush hour period is bi-modal, occurring in the morning and the afternoon, at times that vary by state and urban-rural status. This systematic review and meta-analysis aimed to evaluate the association between the rush hour period and fatal and non-fatal crash injuries. Selected articles were limited to peer-reviewed full-text articles that measured crash injury as an outcome and rush hour as either a predictor, covariate, stratification, or a control variable. A total of 13 articles were identified for systematic review and seven articles were included in the meta-analysis. Across the selected studies, the rush-hour period signified the period of “peak traffic flow.” During the rush hour period, aggressive driving behavior, truck driving, bicycle riding, and precipitation were associated with increased crash events or crash injuries. Across the seven studies included in the meta-analysis, the effective sample size was 220,471. The rush-hour period was associated with a 41% increased risk of fatal crash injury (Pooled RR: 1.41; 95% CI: 1.35 - 1.48). The morning and afternoon rush hour periods were associated with 40% (Pooled RR: 1.40; 95% CI: 1.13-1.67) and 27% (Pooled RR:1.27; 95% CI: 1.10-1.44) increased crash injury risk, respectively. The rush hour period, though less commonly studied as a predictor of fatal and non-fatal crash injuries, represents an important domain in need of crash injury prevention attention. The knowledge of the pattern of crash injuries, as it varies across countries, states, regions, and county can inform policy and intervention, in the presence of competing public health needs.

## 1. Introduction

Road crashes are significant yet preventable causes of morbidity and mortality. Approximately 1.25 million people die each year worldwide from road-related crash injuries (Center for Disease Control and Prevention, 2016). In the United States (U.S.), one person dies every 14 minutes from crash-related events (National Center for Statistics and Analysis, 2019b). While the U.S. recorded its first three-year successive decline in fatal crash counts in over a decade, annual fatal counts in 2018 exceeded 37,000 deaths (National Center for Statistics and Analysis, 2019a). Nevertheless, crash-related morbidity has not substantially declined despite declining fatal crash counts (National Highway Traffic Safety Administration, 2017).

Rush hour represents the period of the day during which the density of humans and vehicles in the road environment is highest. In the U.S., the rush hour period is bi-modal, occurring in the morning and the afternoon (Federal Highway Administration, 2017). The exact period varies across states and rural-urban categorization (Jaffe, 2014). A substantial proportion of fatal and non-fatal crash injuries happen during the rush hour period (HG.org, 2020). For example, in North Carolina, one out of every four crash events occurs during the afternoon rush hour period, and a lesser proportion occurs in the morning rush hour period (Tippett, 2014). Fatal road events account for about an annual average of 300 billion dollars in revenue (Tippett, 2014). The U.S. recorded a decline in road crash injuries death for the first time in 2017 (National Highway Traffic Safety Administration, 2017). While a decrease in fatalities is a measure of success (Injury Prevention Committee, 2017), the ultimate goal is to achieve a zero-traffic death (Ecola, Popper, Silberglitt, & Fraade-Blanar, 2018).

The pattern of crash events during the rush hour or peak periods varies across countries. In the U.S., an urban Florida study reported higher average crash events during the rush hours compared to the non-rush hour period (Shi, Abdel-Aty, & Lee, 2016). A 12-year pooled study using a national database in Spain reported that 51% of all crashes occur during the rush hour period, with injury severity higher in the peak period compared to the non-peak period (Llamazares, Useche, Montoro, & Alonso, 2019). In contrast, a study conducted in an urban city in China reported lower average crash events during the rush hour period compared to the non-rush hour period (Yu, Wang, Yang, & Abdel-Aty, 2016).

Several human and environmental factors are associated with crash events during the rush hour period. A study from an urban center in Florida reported that although the average volume of traffic and traffic volume per lane was less during the rush hour, vehicle occupancy, standard deviation of the vehicular speed, and the congestion index were higher during the rush hour period when compared to non-rush hour period (Shi et al., 2016). Also, six percent of children less than two years of age involved in fatal crash events were unrestrained (Huang, Liu, & Pressley, 2019). Additionally, crash rates among teenage drivers in cities in Virginia were highest between 7 - 8 am and 2 - 6 pm (Vorona et al., 2011). Older drivers with functional impairment are more likely to avoid driving during rush hour compared to those without impairment (Ball et al., 1998).

Despite the vast literature on crash injury etiology, prevalence, and prevention, the rush hour or peak period literature is sparse. It is unknown to what extent fatal and non-fatal crash events are associated with the rush hour period. The presence of a significant relationship between the rush hour period and crash injury risks could present research areas on the unique environmental, behavioral, vehicular, and policy-related factors associated with rush hour-related fatal and non-fatal crash injuries. The rush-hour period may be used as a proxy for intervention aimed at preventing crash injuries if there is data-driven evidence that significant fatal and non-fatal crash injuries occur during this time period. Additionally, the knowledge of the crash injury risk associated with the rush hour may inform policy on lane expansion, utilization of high occupancy vehicle lanes, and road network and design. The research question guiding this literature search is: What is the association of the rush hour period with road crashes? This systematic review and meta-analysis aimed to evaluate the association between the rush hour period and fatal and non-fatal crash injuries.

## 2. Methods

### 2.1. Criteria for selecting peer-reviewed articles

The literature search was limited to peer-reviewed articles published between 2000 and 2020. Selected articles included full-text articles, written in English, with crash injury or event, measured in any form, serving as the outcome measure. Additionally, the rush hour must be measured as either a predictor, covariate, stratification, or control variable. Articles were restricted to the U.S. since traffic characteristics vary widely across countries. Experimental and qualitative studies and studies that do not measure the association between the rush hour period and crash injury or event were excluded.

### 2.2 Search Criteria

The synthesis of literature followed the Preferred Reporting Items for Systematic Reviews and Meta-Analyses (PRISMA) guidelines. Search strings were applied to the advanced search platform of the web pages of the popular databases. Specifically, the researcher searched PsycINFO, PubMed, Academic Search Complete, Cumulative Index of Nursing and Allied Health Literature (CINAHL), Web of Science, Scopus, and Transportation Research International Documentation (TRID). A decision on search adequacy was made when no new item emerged from the databases.

The keywords in all the searches were (((“rush hour”) OR (“peak traffic”) AND ((“road accident”) OR (“road injury”) OR (“road collision”) OR (“road crash”) OR (“traffic accident”) OR (“traffic injury”) OR (“traffic collision”) OR (“traffic crash”) OR (“highway accident”) OR (“highway injury”) OR (”highway crash”) OR (“crash”) OR (“crash injury”))). This search string was applied to the PubMed database and fragmented as multiple searches for all the other databases. Essentially, the multiple searches performed on the other databases used the format “rush-hour” OR “peak traffic” AND the different iterations of “crash injury” stated above. While searching each database, inverted commas, or square or round brackets were applied as appropriate to the database.

Across all the databases, the searches were limited to the inclusion criteria (English, peer-reviewed, 2000 - 2021). The selected articles were imported into an EndNote library. Duplicate articles were removed, and the abstracts were screened for the inclusion and exclusion criteria. Further screening was performed by reading the full texts (Figure 1).

**Figure 1.**
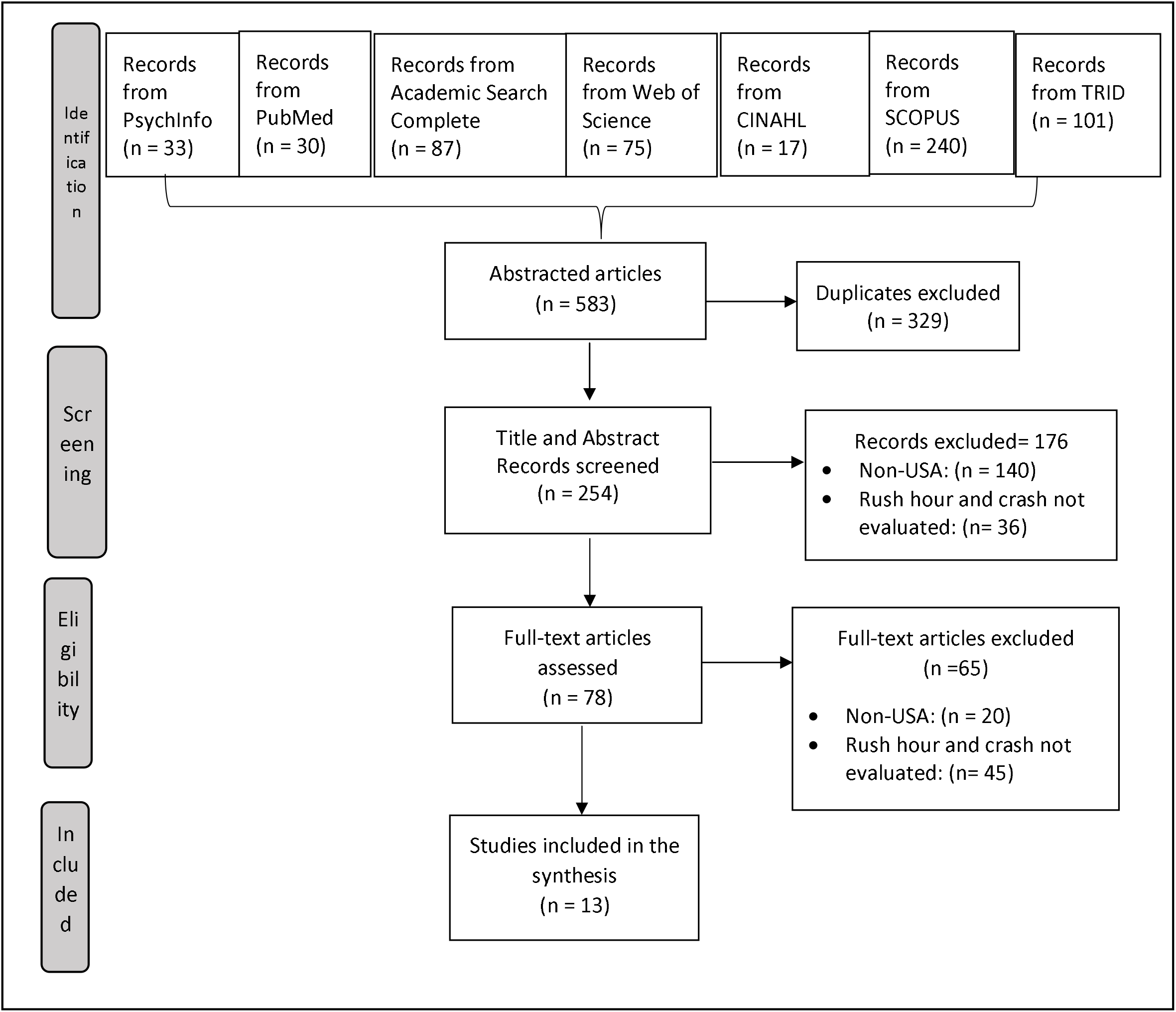
Preferred Reporting Items for Systematic Reviews and Meta-Analyses (PRISMA) flow chart showing the article selection steps for the systematic review

### 2.3. Data selection

A total of 583 hits were generated after the search across the six databases. After excluding 329 duplicate articles, 254 abstracts and titles were screened. Based on the information contained in the title and abstract, 140 non-U.S. articles were excluded and an additional 36 articles were excluded for not meeting the inclusion criteria. After reading through the remaining 78 articles, 65 articles were excluded for not meeting the inclusion and exclusion criteria. A total of 13 articles were selected for synthesis.

### 2.4. Methodological information and analysis

The information gathered from the studies included the study design, data source, study population, study location, sample size, the stated or implied analytical framework, and how the rush hour and crash injury or event were defined. The coding scheme was designed to capture the inferential statistics used, and the odds or risks of crash injury or event associated with the rush hour period were documented.

The data reporting methods across the papers were streamlined for readability and consistency. The confidence intervals of coefficients or odds ratio were reported in preference to standard errors. For papers that reported only the coefficients and standard errors, the confidence intervals were computed using the formula: *CI = X ± 1.96 SE* where CI represents the lower and upper confidence intervals, X represents the coefficient, and SE represents the standard error. Logit coefficients were converted to odds ratio using the formula: *OR = e^x^* where OR represents the odds ratio, e represents the exponential function, and X represents the coefficient.

### 2.5. Quality assessment

Methodological quality assessment was performed using the Downs and Black checklist (Downs & Black, 1998). The checklist is a 27-item list that captures the reporting, external, and internal validity measures of the study and the power of the study. The Downs and Black checklist has high reliability and validity values (Downs & Black, 1998). The maximum score is 25 for non-randomized studies and 28 for randomized studies. Similar to previous studies, scores of 26 and higher were determined as excellent, 20 – 25 as good, 15 to 19 as fair, and 14 and lower as poor (Hooper, Jutai, Strong, & Russell-Minda, 2008).

### 2.6. Meta-Analysis

A subset of the selected papers with reported odds, risks, or estimates of crash injury associated with the rush hour period was meta-analyzed. While some studies reported a single estimate for crash events during specified rush hour period, other provided estimates for each hour. Studies that did not report the odds ratio or coefficients and the associated confidence intervals were excluded. The effect sizes were computed for all selected studies. Subgroup analysis was performed by assessing the differences in the effect sizes in the morning and afternoon rush hour periods, and by fatal and non-fatal crash injuries. A meta-regression analysis was performed using the rush hour periods as predictors.

Since the data on crash injury or events were drawn from heterogeneous sources, a random effect was used in measuring the effect size of rush hour. Essentially, the random effect is stated thus: 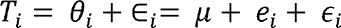, where *T_i_* represents the observed effect, *θ_i_* represents the true effect, which is a sum of the mean (*μ*) and the within-study error (*e_i_*), and *∈_i_* represents the between study error (Borenstein, Hedges, Higgins, & Rothstein, 2010). For each study, weights were applied. Weights 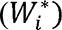 were calculated using the inverse variance 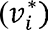, a nuanced measure of the sample size, using the formula: 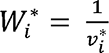. The weighted mean (*T*), therefore, represents the sum of the products of effect size (*T_i_*) and the weights (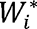) divided by the sum of the weights 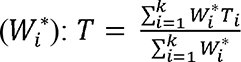 (Borenstein et al., 2010).

Risk ratios were used in the meta-analysis to measure the effect size since most of the selected articles reported the odds and risk ratios of crash injury. The pooled risk ratio (RR) was assessed using the Mantel-Haenszel method using the formula: 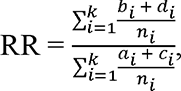 where a, b, c, and d represent exposed cases, non-exposed cases, exposed non-cases, non-exposed non-cases, respectively, and n is the sum of a, b, c, and d (Schmidt & Kohlmann, 2008). Standard errors (SE) were estimated using the formula: SE (ln(RR)) = 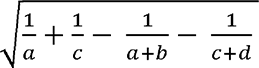 and the 95% confidence intervals were estimated from the standard errors using the formula: 95% CI = exp (ln(RR) ± 1.96 x SE (ln(RR))) (Schmidt & Kohlmann, 2008).

Coefficient values, calculated using the risk estimates’ natural log transformation, were used to assess publication bias. Test of heterogeneity (I^2^), which reports the variability in the result, were reported. I^2^ represents the percentage of error due to between-study variations. Values of less than 25% indicate excellent, while values between 25 and 50% represent moderate heterogeneity (Higgins, Thompson, Deeks, & Altman, 2003). Values above 50% suggest some degree of homogeneity in the studies (Higgins et al., 2003). Funnel plots, trim and fill analysis, and Egger’s regression test was used to assess publication bias. With the funnel plot, the symmetrical location of the studies around the population effect size suggests no bias (Peters, Sutton, Jones, Abrams, & Rushton, 2008). The Egger test is a quantitative measure of the publication bias, with a p-value less than 0.05 suggestive of publication bias (Lin & Chu, 2018). Using the trim and fill analysis, studies were trimmed to make the effect size less affected by publication bias and studies are imputed to achieve symmetrical distribution of the studies (Shi & Lin, 2019). Statistical analysis was performed using STATA version 16 (StataCorp, 2020).

## 3. Results

### 3.1 Study characteristics

Of the 583 articles identified using the search criteria, 13 papers met the inclusion and exclusion criteria, and these studies were included in the synthesis. All the selected papers used retrospective designs. Secondary data sources used in the selected studies ranged widely from national census data such as the Fatality Analysis Reporting System (FARS) (Huang, Liu, & Pressley, 2019; Stevens, Schreck, Saha, Bell, & Kunkel, 2019) and the National Motor Vehicular Crash Causation Survey (NMVCCS) (Paleti, Eluru, & Bhat, 2010) to state-level data (Call, Medina, & Black, 2019; Cook, Knight, & Olson, 2005; Das, Pande, Abdel-Aty, & Santos, 2008; Duddu, Kukkapalli, & Pulugurtha, 2019; Haleem & Gan, 2013; Jang, Chung, Ragland, Chan, & Transportation Research, 2008; Kim, Kim, Ulfarsson, & Porrello, 2007; Lee, Dittberner, & Sripathi, 2007) (Table 1). The purpose of the papers varied widely. While some papers assessed the natural and environmental determinants of crash events or injuries (Call et al., 2019; Das et al., 2008; Jang et al., 2008; Lee et al., 2007; Marquis & Wang, 2015; Stevens et al., 2019), others assessed the association of risky driving behaviors and the occurrence of crash events or injuries (Cook et al., 2005; Duddu et al., 2019; Huang et al., 2019; Ma, Hao, Xiang, & Yan, 2018; Paleti et al., 2010). The study population focused on either the individuals involved in the crash (Huang et al., 2019; Kim et al., 2007; Paleti et al., 2010) or crash events (Call et al., 2019; Cook et al., 2005; Das et al., 2008; Jang et al., 2008; Lee et al., 2007; Ma et al., 2018; Marquis & Wang, 2015). Also, the sample sizes had a wide range - from approximately 1,000 (Marquis & Wang, 2015) to over 620,000 (Cook et al., 2005). Using the Down and Black checklist, the qualities of the studies were low to average.

**Table 1:** Characteristics of the papers selected for the systematic review

| Authors (Year) | Purpose | Study Design | Data Source | Study Population | Sample Size | Quality Scores* (/28) |
| --- | --- | --- | --- | --- | --- | --- |
| Call et al., (2019) | Assessed how crashes related to weather differed by severity, date and time, driver, and other environmental conditions | Retrospective | 2012-2016 Utah DOT | Motor vehicular crash events | 14,863 | 8 |
| Cook et al., (2005) | Determined the characteristics and outcomes associated with aggressive driving crashes compared to DUI and non-DUI non-aggressive crashes | Retrospective | 1992-2003 Utah DOT<br>1996-2002 ED, University of Utah<br>1992-2002 DOH, University of Utah | Motor vehicular crash events | 621,451 | 8 |
| Das et al., (2008) | Assessed the factors associated with crashes and crash injury severity related to intersections on multilane arterial roads | Retrospective | 2002 – 2005 crash data from Florida DOT | Motor vehicular crash events | 1,575 | 12 |
| Duddu et al., (2019) | Assessed the factors associated with crash injury severity among teen drivers | Retrospective | 2011 – 2013 Highway Safety Information System, N.C. | Crashes among teen drivers (15 – 19 years) | 62,990 | 13 |
| Haleem et al., (2013) | Assessed the predictors of injury severity on urban freeways | Retrospective | 2001 – 2005 crash data from Florida DOT | Non-fatal crash injury events | 56,727 | 13 |
| Huang et al., (2019) | Assessed the characteristics of fatal crashes among children with missing restraint documentation | Retrospective | 2008 – 2015 FARS data from NHTSA | Child crash injury severity | 4,966 | 13 |
| Jang et al., (2008) | Compared crash events on different traffic routes | Retrospective | 1999 – 2003 crash data from California DOT | Motor vehicular crash events | 6,748 | 8 |
| Kim et al., (2007) | Predicted the occurrence of bicycle-motor crashes using human, vehicular, land, environmental, and temporal characteristics | Retrospective | 1997 – 2002 N.C. police-reported data | Single motorist and bicyclist road crash | 2934 | 13 |
| Lee et al. (2007) | Assessed the factors associated with lane-specific traffic safety | Retrospective | 2002 – 2004 Washington DC and Virginia police crash report | Motor vehicular crash events | 1,613 | 13 |
| Ma et al., (2018) | Assessed the factors associated with crash injury severity among drivers with and without aggressive driving behaviors | Retrospective | 2005 – 2014 Federal Railroad Administration database | Highway-rail crash events | 25,945 | 13 |
| Marquis et al., (2015) | Assessed the temporal effects of truck crash events | Retrospective | 2008 – 2011 crash data from New York State DOT | Motor vehicular crash events | 1,086 | 13 |
| Paleti et al., (2010) | Assessed the relationship of aggressive driving behavior and road accident injury, and the moderating effect of environmental and temporal characteristics | Retrospective | 2005 – 2007 NMVCCS data | Crash observations of drivers | 2,315 | 14 |
| Stevens et al., (2019) | Assessed the association of precipitation and precipitation intensity on fatal road crashes and how the association changes across regions and the times of the day | Retrospective | 2006 – 2011 FARS data | Fatal crash events | 125,012 | 13 |

DOT: Department of Transportation; ED: Emergency Department; DoH: Department of Health; NC: North Carolina; DUI: Driving Under the Influence of alcohol and drugs; CAS: Crash Analysis System; FARS: Fatality Analysis Reporting System; NHTSA: National Highway Traffic Safety Administration; NMVCCS: National Motor Vehicular Crash Causation Survey; \*Quality scores assessed using the Downs and Black checklist

### 3.2 Definition of the Rush Hour Period

Across the selected studies, the rush hour period was referred to as the period with “peak traffic volume” (Call et al., 2019; Cook et al., 2005; Das et al., 2008; Duddu et al., 2019; Haleem & Gan, 2013; Huang et al., 2019; Jang et al., 2008; Kim et al., 2007; Ma et al., 2018; Marquis & Wang, 2015; Paleti et al., 2010) or the period with the highest road crash events (Stevens et al., 2019) (Table 2). The rush-hour period defined the time during which certain road processes were activated such as the period when the road shoulders are opened to accommodate increased traffic flow (Lee et al., 2007). The rush-hour period was not defined uniformly across the studies. The morning rush hour period started as early as 5 or 5:30 am in densely populated regions like Washington DC and Virginia (Lee et al., 2007) or states with a large population size like California (Jang et al., 2008). Additionally, studies conducted in densely populated areas like New York and Florida reported that the morning rush hour period ended at either 10 or 11 am (Haleem & Gan, 2013; Marquis & Wang, 2015). The median morning rush hour period starts at 6 am and ends at 9 am. The duration of the morning rush hour period hours was as short as one hour (Call et al., 2019) and as long as four hours (Haleem & Gan, 2013; Jang et al., 2008; Kim et al., 2007; Marquis & Wang, 2015). Similarly, the duration of the afternoon rush hour period was as short as two hours (Call et al., 2019; Cook et al., 2005; Ma et al., 2018) and as long as four hours (Haleem & Gan, 2013; Jang et al., 2008; Lee et al., 2007; Paleti et al., 2010). Across the studies, the median afternoon rush hour period started at 3 pm and ended at 7 pm. Among the selected studies, rush hour served different roles in research. Some studies used the rush hour period as a predictor variable (Das et al., 2008; Duddu et al., 2019; Haleem & Gan, 2013; Huang et al., 2019; Kim et al., 2007; Lee et al., 2007; Ma et al., 2018; Stevens et al., 2019), as an interaction or stratification variable (Marquis & Wang, 2015; Paleti et al., 2010), or only for descriptive purposes (Call et al., 2019; Cook et al., 2005).

**Table 2:** Rush hour definitions and statistical conceptualization

| Authors (Year) | Conceptual definition of rush hour | Reference time |  | State Location | Usage | Statistical Measure |
| --- | --- | --- | --- | --- | --- | --- |
|  |  | AM | PM |  |  |  |
| Call et al., (2019) | Time of peak traffic | 7 – 9 | 5 – 7 | Utah | Descriptive | Proportion |
| Cook et al., (2005) | Time of peak traffic | 7 – 8 | 4 - 6 | Utah | Descriptive | Proportion |
| Das et al., (2008) | Time of peak traffic | 7 – 9:30 | 4 - 7 | Florida | Predictor | Bivariate Probit<br>Ordinal Probit |
| Duddu et al., (2019) | Time of peak traffic | 6 – 9 | 3 – 6 | North Carolina | Predictor | Partially Proportional<br>Ordinal Logistic |
| Haleem et al., (2013) | Time of peak traffic | 7 - 11 | 3 – 7 | Florida | Predictor | Mixed Logit<br>Binary Logit |
| Huang et al., (2019) | Time of peak traffic | 7 – 9:30 | 3:30 – 6 | All U.S. States | Predictor | Logistic Regression |
| Jang et al., (2008) | Time of peak traffic | 5 – 9 | 3 – 7 | California | Stratified<br>outcome<br>measure | Proportion |
| Kim et al., (2007) | Time of peak traffic | 6 – 10 | 3 – 6 | North Carolina | Predictor | Multinomial Logit |
| Lee et al. (2007) | The period when the road shoulders are opened for traffic flow | 5:30 – 9:30 | 3 – 7 | Washington DC;<br>Virginia | Predictor | Negative Binomial<br>Regression |
| Ma et al., (2018) | Time of peak traffic | 6 – 9 | 4 – 6 | All U.S. States | Predictor | Mixed Logit |
| Marquis et al., (2015) | Time of peak traffic | 6 – 10 | 3 – 7 | New York | Stratifying<br>variable | Proportion |
| Paleti et al., (2010) | Time of peak traffic | 6 – 9 | 3 – 7 | Texas | Interaction<br>variable | Ordinal logistic |
| Stevens et al., (2019) | Period with the highest crash event in the day | 6 – 9* | 3 – 6* | All U.S. States | Predictor | Relative Risk |

\*Not stated but information obtained from the National Highway Traffic Safety Administration publication

### 3.3 Road Crash Definition

Crash, as an outcome, was measured as either crash event (Call et al., 2019; Cook et al., 2005; Jang et al., 2008; Lee et al., 2007; Marquis & Wang, 2015) or crash injury (Das et al., 2008; Duddu et al., 2019; Haleem & Gan, 2013; Huang et al., 2019; Kim et al., 2007; Ma et al., 2018; Paleti et al., 2010; Stevens et al., 2019) across the selected studies (Table 3). Crash events were defined numerically as counts of crashes (Call et al., 2019; Cook et al., 2005; Jang et al., 2008; Lee et al., 2007; Marquis & Wang, 2015) or categorically as the occurrence of a crash event (Das et al., 2008). Crash injuries were defined as either binary or as an ordinal variable. Binary definition of crash injury was either defined as fatal or non-fatal or severe or non-severe nonfatal injury (Haleem & Gan, 2013). Ordinal definition of crash injury used either a three-point or five-point scale (Das et al., 2008; Duddu et al., 2019; Huang et al., 2019; Kim et al., 2007; Ma et al., 2018; Paleti et al., 2010). The three-point scale was defined as killed/fatal, injured, and property damage only/no injury (Duddu et al., 2019; Huang et al., 2019; Ma et al., 2018) while the five-point scale was defined as fatal, incapacitating injury, non-incapacitating injury, possible injury, and no injury/property damage only (Das et al., 2008; Kim et al., 2007; Paleti et al., 2010). Across the selected studies, the analytical approaches involved the use of proportions, bivariate logit/ logistic regression, or ordinal probit, logit, or logistic regression models, multinomial logit regression model, mixed logit models, and negative binomial regression models (Table 2).

**Table 3:**
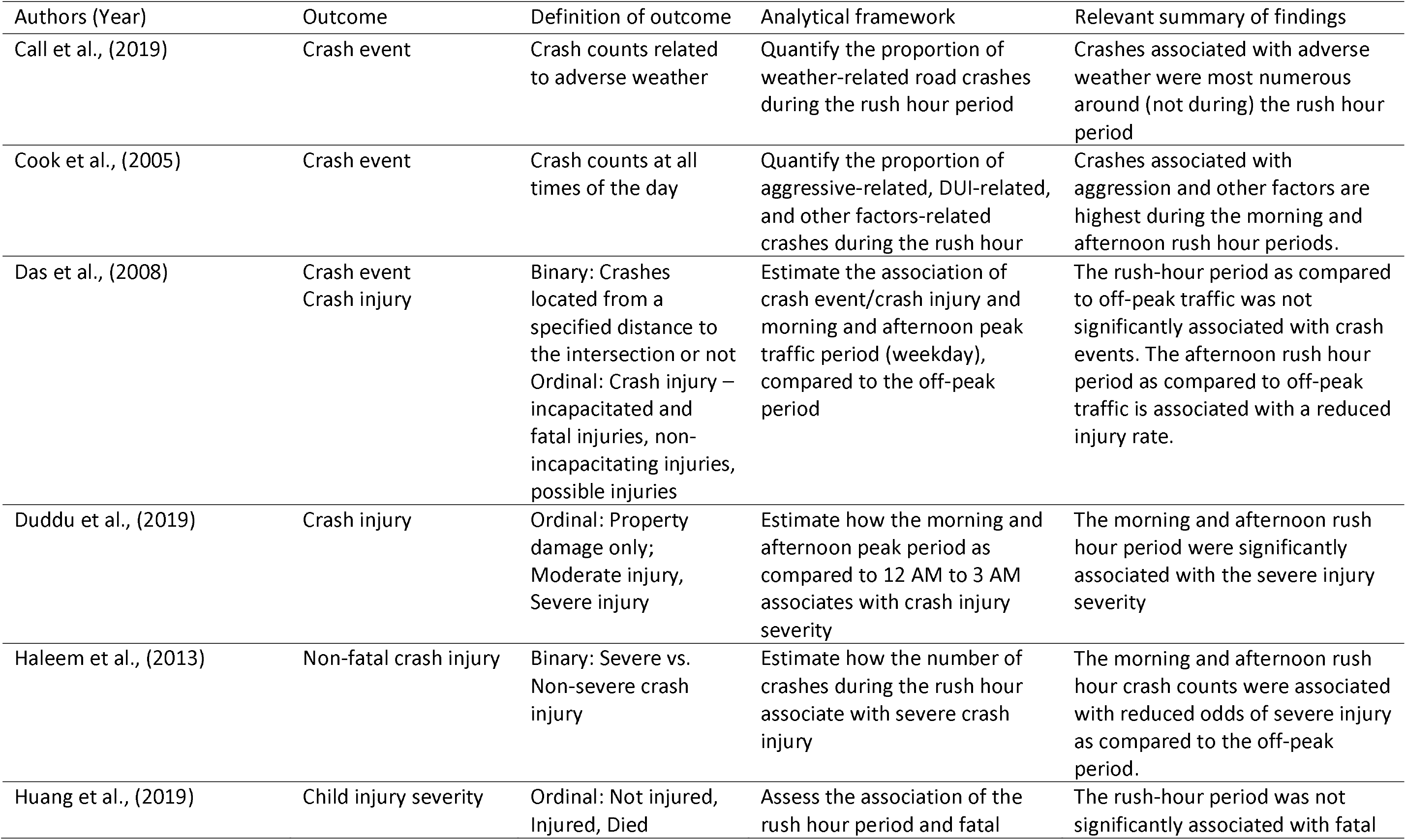
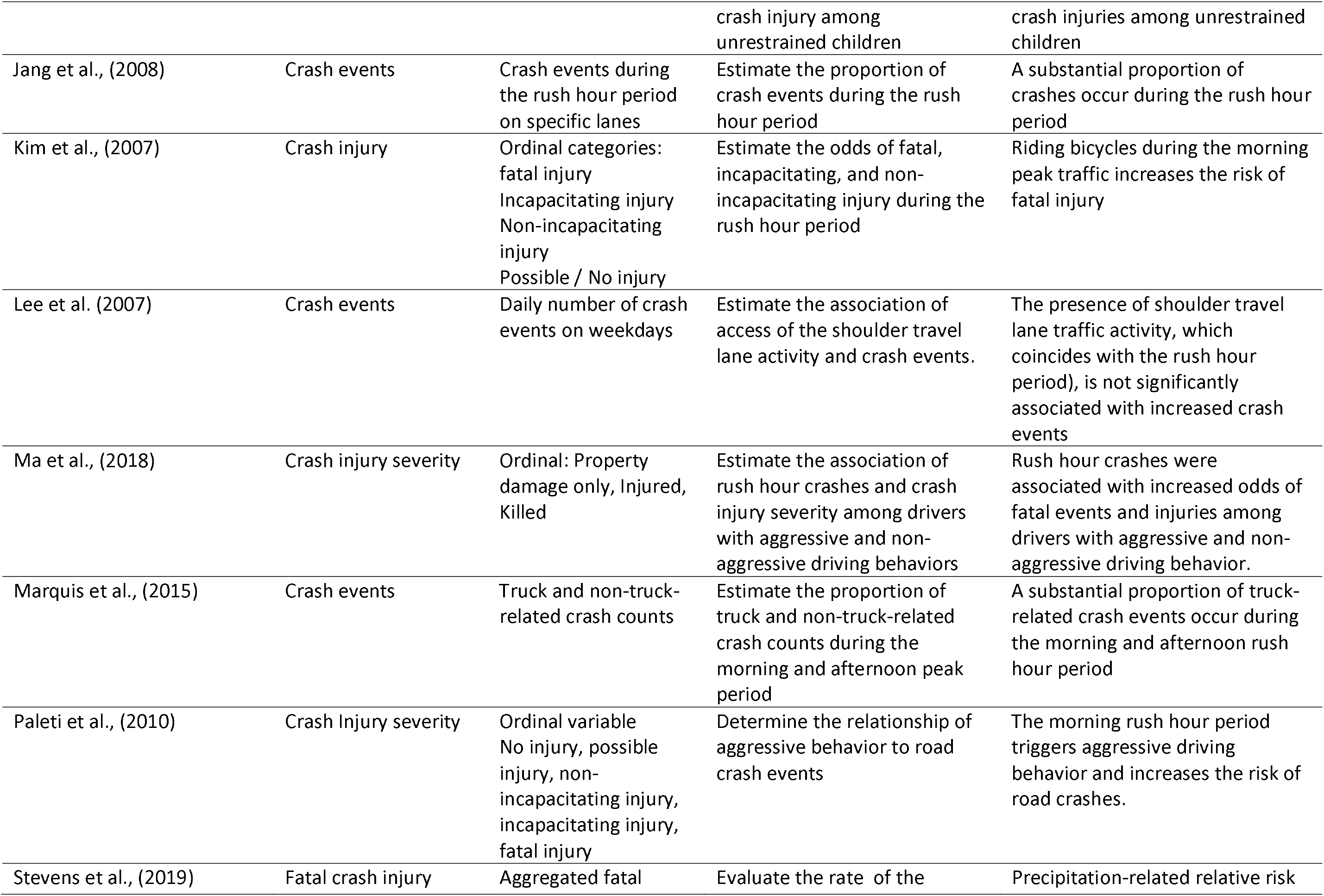
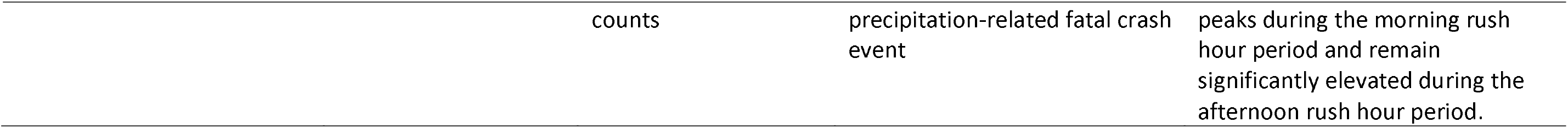
Summary of the how road crash was defined and measured, as well as the rush hour-related analytical framework and summary findings in the selected studies.

A substantial proportion of crash events occur during the rush hour period (Jang et al., 2008; Marquis & Wang, 2015). There were conflicting results on the relationship of the rush hour study was crash events or crash injury. A few studies reported that the rush hour period was significantly associated with fatal or severe injury severity (Duddu et al., 2019; Huang et al., 2019; Ma et al., 2018) while others reported that the rush hour was associated with reduced odds of crash injury severity (Haleem & Gan, 2013), reduced injury rates (Das et al., 2008), or reduced fatal injuries (Huang et al., 2019).

Across the selected studies, individual, vehicular, and environmental determinants of crash injury or events were assessed. Within the domain of the individual determinants of crash injury, the selected studies reported that the rush hour period was a trigger for aggressive driving behavior, with such behaviors more in the morning than in the afternoon rush hour period (Paleti et al., 2010). Aggressive driving behavior-related crashes were more during the rush hour period (Cook et al., 2005) compared to other times of the day.

Bicycle riding and truck driving were the vehicle determinants that emerged from the selected studies. Bicycle riding during the morning rush hour period was associated with an increased risk of fatal injury (Kim et al., 2007) while truck-related crash events were more in the morning rush hour period compared to the afternoon rush hour period (Marquis & Wang, 2015). Within the domain of the built and natural road environment, the rush hour period was associated with increased intersection-related crash rates (Das et al., 2008). Also, adverse weather-related crashes were more during the rush hour period (Call et al., 2019) and rates of precipitation-related fatal injury were significantly elevated during the morning and afternoon rush hour periods (Stevens et al., 2019)). The presence of shoulder travel lane, activated during the rush hour period, was not associated with significantly increased or reduced rates of fatal crash injuries (Lee et al., 2007)

### 3.4 Prevalence of Crash Injuries During the Rush Hour

About 23% and 21% of truck crashes occur during the morning and afternoon rush hour periods (Marquis & Wang, 2015) (Table 4). Also, about 9% and 33% of bicycle-related crashes occur during the morning and afternoon rush hour periods, respectively (Kim et al., 2007). Also, 6.5% and 8.5% of aggressive driving-related crash events occur during the morning and afternoon rush hour periods, respectively (Cook et al., 2005). The prevalence of crash events due to driving under the influence of alcohol or substance use (DUI) was 2% and 8.5% during the morning and afternoon rush hour periods, respectively (Cook et al., 2005). Across the selected studies, 14 – 16% and 7 – 11% of weather-related crashes occur during the morning and afternoon rush hour periods, respectively (Call et al., 2019).

**Table 4:**
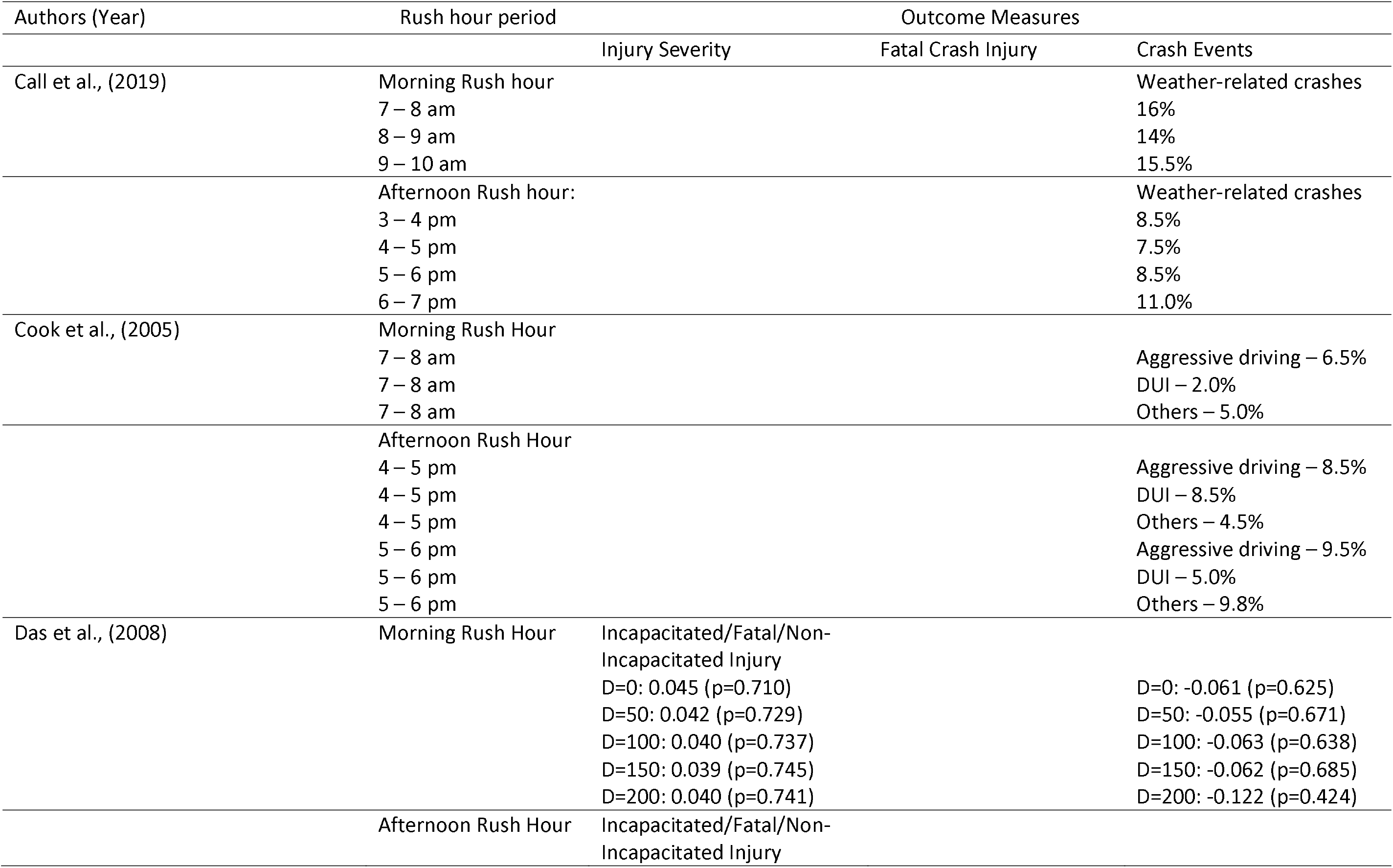
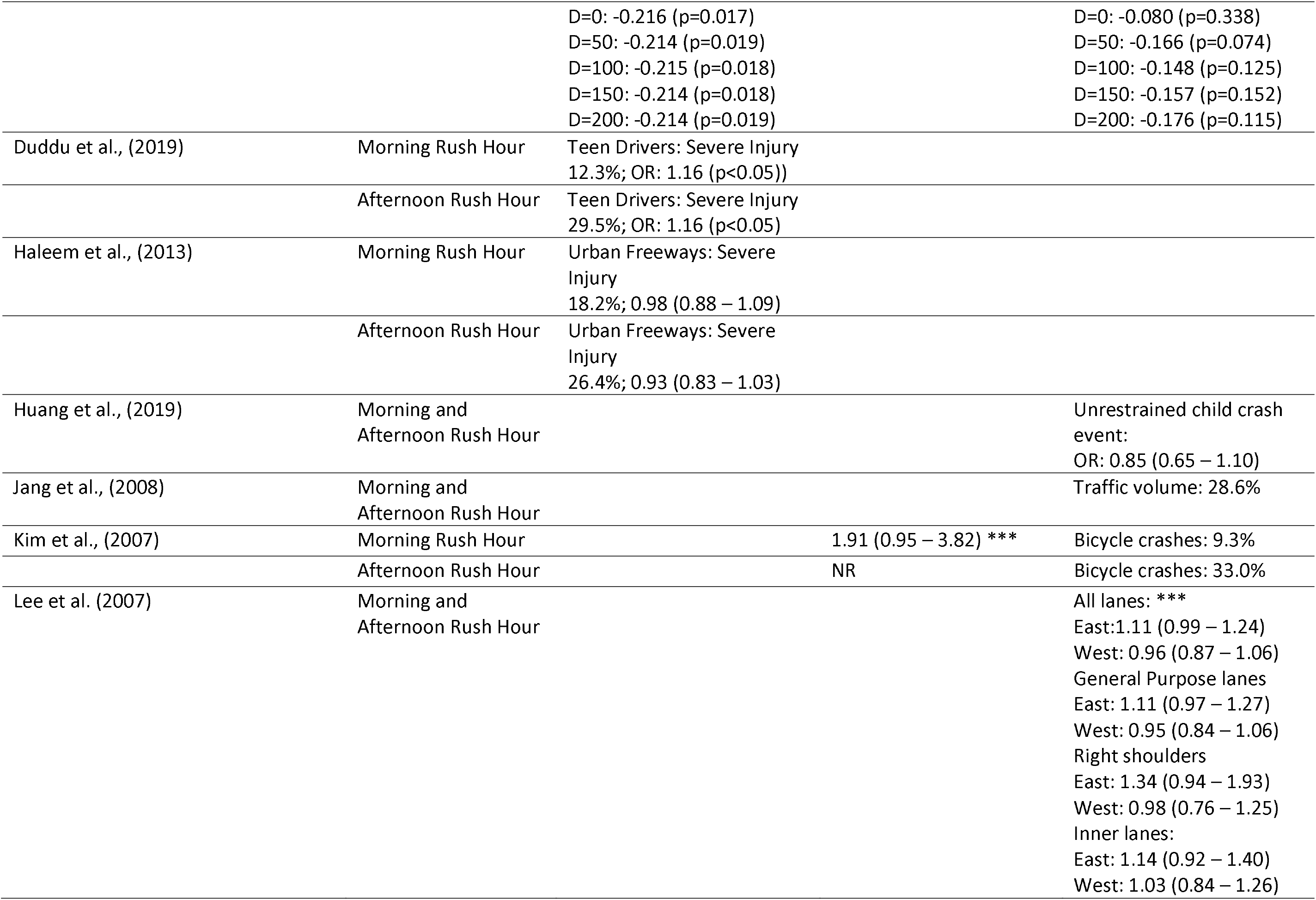
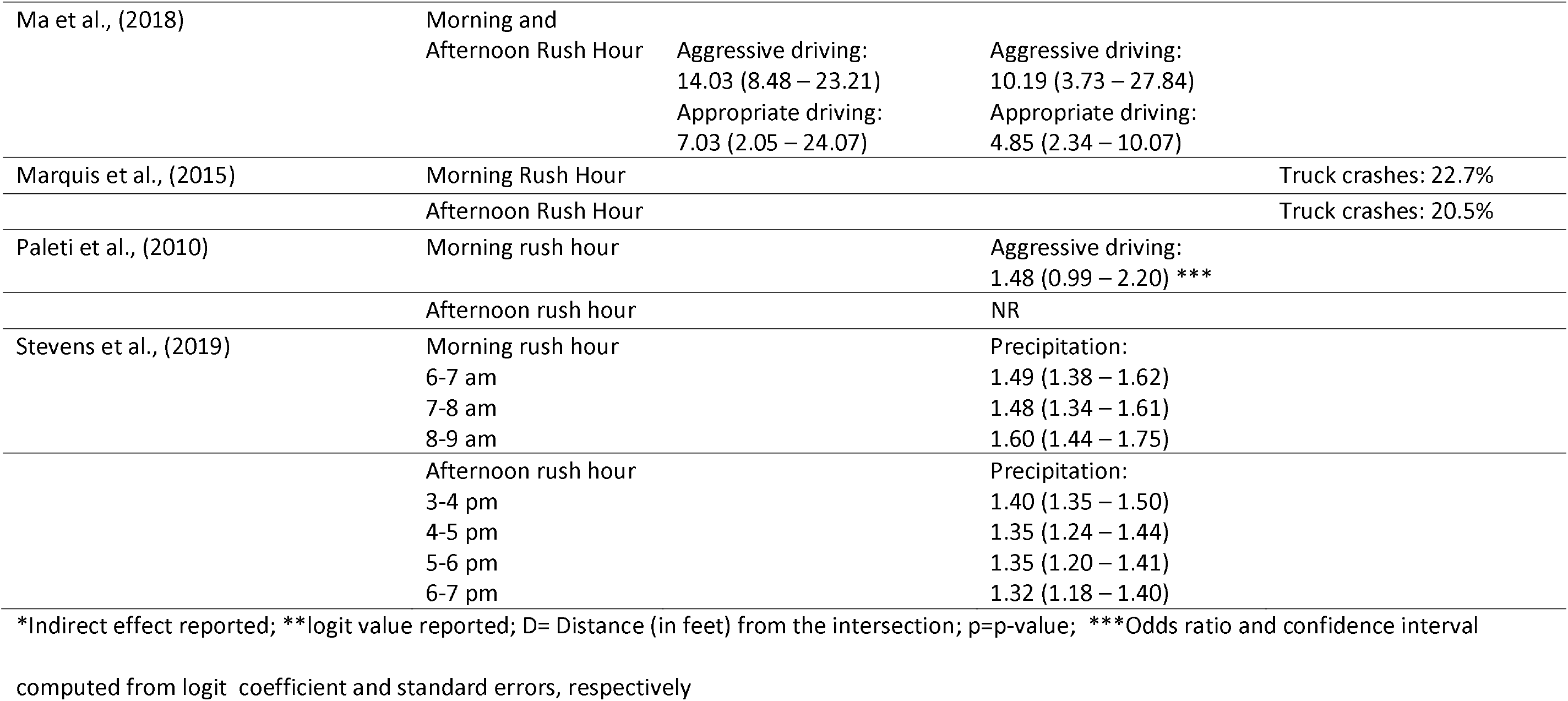
Summary of the relationships reported across the studies between rush hour period and road crash injuries

### 3.5 Rush Hour and Crash Injuries

During the rush hour period, precipitation was significantly associated with increased fatal crash injury risk (Stevens et al., 2019) (Table 4). There was no statistical relationship between intersection-related severe and fatal crash injury and the morning rush hour traffic volume (Das et al., 2008). However, the afternoon rush hour traffic volume was associated with reduced odds of intersection-related fatal and severe injury severity (Das et al., 2008). The rush-hour period was significantly associated with increased odds of severe injury among teen drivers (Duddu et al., 2019). Aggressive driving behavior was associated with increased odds of fatal crash injuries and non-fatal injuries during the rush hour period (Ma et al., 2018). Also, the rush hour period was associated with increased fatal injury among unrestrained children (Huang et al., 2019).

### 3.6 Summary of Meta-analysis

A total of seven studies with reported risks, odds, or coefficients of fatal and non-fatal crash injury from the rush hour period were selected for the meta-analysis (Table 5). The effective sample size, excluding duplicated studies, was 220,471. The seven studies provided 24 point estimates for either or both morning and afternoon rush hour periods. Less than half of the selected studies reported separate crash injury estimates for the morning and afternoon rush hour period (Haleem & Gan, 2013; Kim et al., 2007; Stevens et al., 2019). Stevens et al. (2019) reported crash risks for every hour during the rush hour period while Lee et al. (2007) reported estimates for different lanes during the rush hour period. Ma et al. (2018) reported the odds of severe injury due to aggressive and appropriate driving during the rush hour period.

**Table 5:** Summary of studies used in the meta-analysis

| Study | Sample Size* | Crash Injury | Period | Effect Size (95% CI) | % Weight |
| --- | --- | --- | --- | --- | --- |
| Haleem et al., 2013 | 56727 | Non-fatal | AM | 0.98 (0.88 - 1.09) | 6.05 |
| Haleem et al., 2013 | 56727 | Non-fatal | PM | 0.93 (0.83 - 1.03) | 6.08 |
| Huang et al., 2019 | 4966 | Fatal | BOTH | 0.85 (0.05 - 1.16) | 2.31 |
| Kim et al., 2007 | 2934 | Fatal | AM | 1.91 (0.95 - 3.82) | 0.5 |
| Lee et al., 2007 | 559 | Crash | BOTH | 1.11 (0.99 - 1.24) | 5.9 |
| Lee et al., 2007 | 643 | Crash | BOTH | 0.96 (0.87 - 1.06) | 6.12 |
| Lee et al., 2007 | 175 | Crash | BOTH | 1.14 (0.92 - 1.4) | 4.82 |
| Lee et al., 2007 | 128 | Crash | BOTH | 1.03 (0.84 - 1.26) | 5.12 |
| Lee et al., 2007 | 379 | Crash | BOTH | 1.11 (0.97 - 1.27) | 5.69 |
| Lee et al., 2007 | 507 | Crash | BOTH | 0.95 (0.84 - 1.06) | 6.01 |
| Lee et al., 2007 | 90 | Crash | BOTH | 1.34 (0.94 - 1.93) | 2.66 |
| Lee et al., 2007 | 91 | Crash | BOTH | 0.98 (0.76 - 1.25) | 4.77 |
| Ma et al., 2018 | 11481 | Fatal | BOTH | 14.03 (8.48 - 23.21) | 0.02 |
| Ma et al., 2018 | 11481 | Fatal | BOTH | 7.03 (2.05 - 24.07) | 0.01 |
| Ma et al., 2018 | 14464 | Non-fatal | BOTH | 4.85 (2.34 - 10.07) | 0.07 |
| Ma et al., 2018 | 14464 | Non-fatal | BOTH | 10.19 (3.73 - 27.84) | 0.01 |
| Paleti et al., 2010 | 2315 | Fatal | BOTH | 1.48 (0.99 - 2.20) | 2.06 |
| Stevens et al., 2019 | 125012 | Fatal | AM | 1.49 (1.38 - 1.62) | 5.94 |
| Stevens et al., 2019 | 125012 | Fatal | AM | 1.48 (1.34 - 1.61) | 5.82 |
| Stevens et al., 2019 | 125012 | Fatal | AM | 1.6 (1.44 - 1.75) | 5.65 |
| Stevens et al., 2019 | 125012 | Fatal | PM | 1.4 (1.35 - 1.50) | 6.23 |
| Stevens et al., 2019 | 125012 | Fatal | PM | 1.35 (1.24 - 1.44) | 6.08 |
| Stevens et al., 2019 | 125012 | Fatal | PM | 1.35 (1.20 - 1.41) | 6.05 |
| Stevens et al., 2019 | 125012 | Fatal | PM | 1.32 (1.18 - 1.40) | 6.01 |

\*Unique sample size=220,471 excluding the duplicated studies that reported morning and afternoon rush hour estimates

### 3.7 Meta-analysis: Rush Hour and Crash Event/Injury

Across all the seven selected studies and 24 point estimates, the pooled risk for rush hour as a predictor of crash injury was 1.21 (95% CI: 1.11 - 1.32) (Figure 2A). The test for heterogeneity was considerably high across all selected studies since some studies had multiple entries (I^2^ = 89.02%). There was visual evidence of publication bias, with most of the selected studies asymmetrically located on one side the pooled effect size (Figure 2B). The Egger test was statistically significant (p<0.001), suggesting that there was statistical evidence of publication bias (Table 6). Using the Trim and Fill analysis, five studies were imputed (n=29) (Figure 2C). The effect size, after adjusting for publication bias, was 1.20 (95% CI: 1.10 – 1.31), suggesting that the rush hour period was associated with a 20% increased risk of crash injury/event (Table 6).

**Figure 2:**
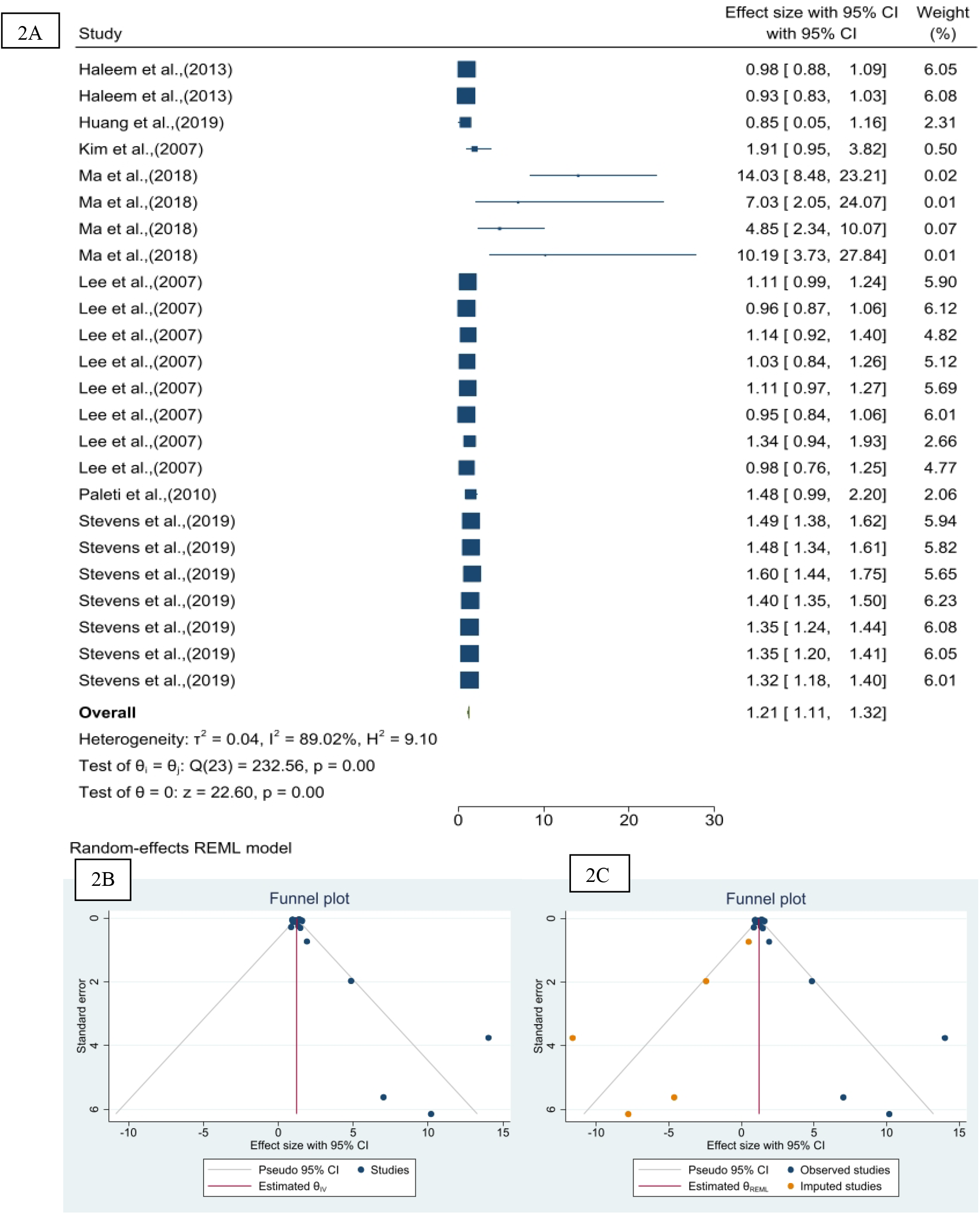
Forest plot (A), funnel plot (B), and Trim and Fill analysis plot (C) of the selected studies showing the effect size and publication bias and the adjustment for publication bias

**Table 6:** Summary of the Trim and Fill analysis showing the adjusted effect size and the Egger test evaluation of publication bias across the selected studies.

| Trim and Fill Analysis |  |  |
| --- | --- | --- |
| Studies | Theta | 95% CI |
| Observed (n=24) | 1.21 | 1.11 – 1.32 |
| Observed + Imputed (29) | 1.20 | 1.10 – 1.31 |
| Regression-based Egger test for small-study effects |  |  |
| Beta (Standard Error) | Z-value | p-value |
| 1.44 (0.42) | 3.43 | 0.001 |

### 3.8 Sub-group Meta-Analysis: Fatal and Non-Fatal Crash Injury

Six studies measured crash outcomes as either fatal or non-fatal crash injuries. A total of 16 point estimates emerged from these six studies (Figure 3A). The rush-hour period was associated with a 32% increased risk of fatal and non-fatal crash injury (Pooled Risk Ratio: 1.32; 95% CI: 1.18 – 1.46). The test for heterogeneity was considerably high across all selected studies since some studies had multiple entries (I^2^ = 89.59%). There was visual evidence of publication bias, with most of the selected studies asymmetrically located on one side the pooled effect size (Figure 3B). The Egger test was statistically significant (p<0.001), suggesting that there was statistical evidence of publication bias (Table 7). Using the Trim and Fill analysis, five studies were imputed (n=21) (Figure 2C). The effect size, after adjusting for publication bias, was 1.31 (95% CI: 1.17 – 1.45), suggesting that the rush hour period was associated with 31% increased fatal and non-fatal crash injury (Table 7).

**Figure 3:**
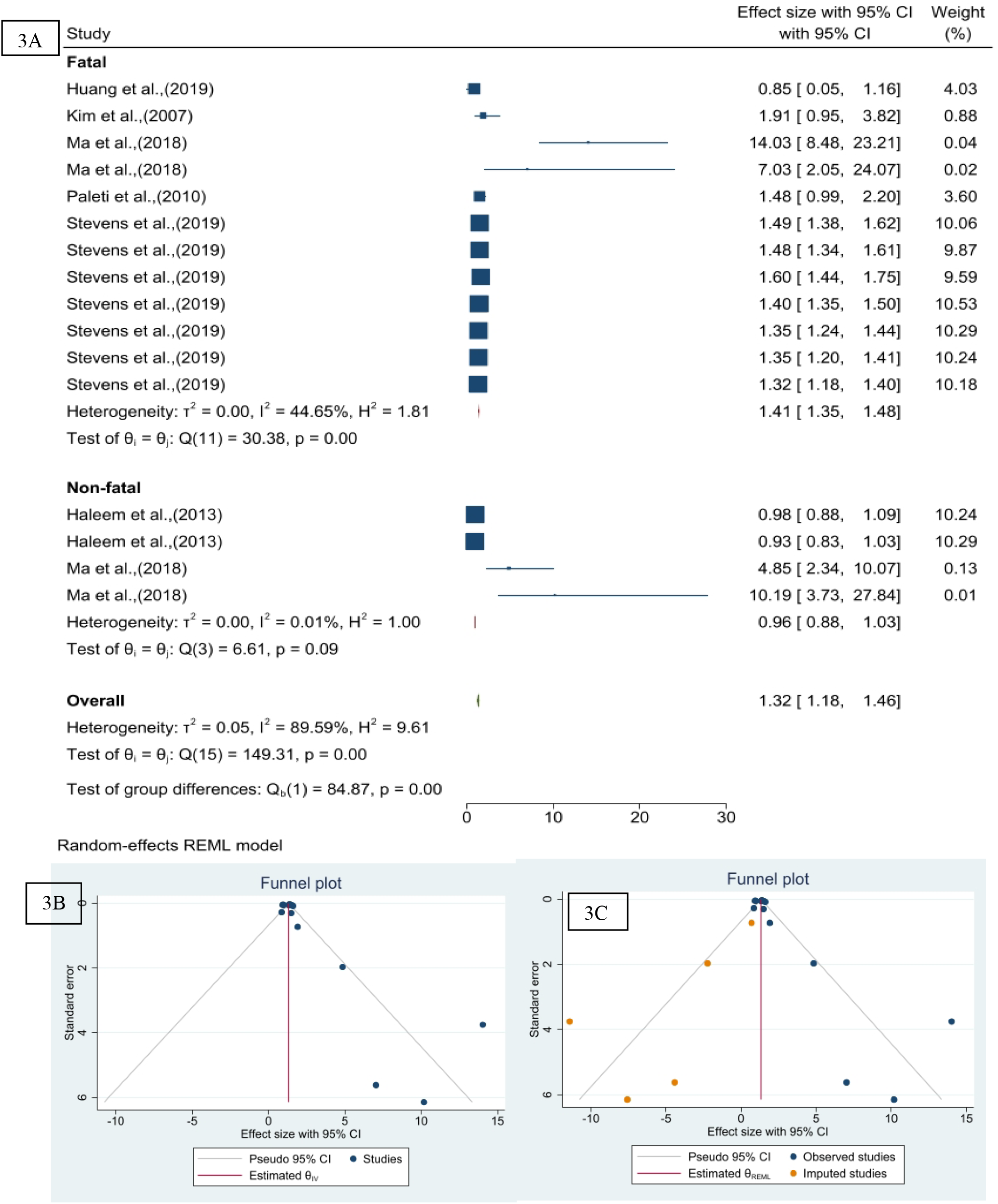
Forest plot (A) showing the sub-group analysis by fatal and non-fatal status, funnel plot (B) visualizing publication bias, and Trim and Fill analysis plot (C) showing the adjustment for publication bias

**Table 7:** Summary of the Trim and Fill analysis of the sub-group analysis by fatal and non-fatal crash injury, showing the adjusted effect size and the Egger test evaluation of publication bias across the selected studies.

| Trim and Fill Analysis |  |  |
| --- | --- | --- |
| Studies | Theta | 95% CI |
| Observed (n=16) | 1.32 | 1.18 – 1.46 |
| Observed + Imputed (21) | 1.31 | 1.17 – 1.45 |

| Regression-based Egger test for small-study effects |  |  |
| --- | --- | --- |
| Beta (Standard Error) | Z-value | p-value |
| 1.47 (0.435) | 3.38 | 0.001 |

The subgroup analysis, measured by fatal and non-fatal crash injury status showed that the rush hour was associated with a 41% increased risk of fatal crash injury (Pooled Risk Ratio: 1.41; 95% CI: 1.35 – 1.48). There was moderate heterogeneity across the selected studies (I^2^ = 44.65%). The rush-hour period was associated with a 4% reduced risk of non-fatal crash injury, but the association was not statistically significant (Pooled Risk Ratio: 0.96; 95% CI: 0.88 – 1.03). There was excellent heterogeneity across the studies (I^2^ = 0.01%). The funnel plot of the subgroup analysis (fatal and non-fatal crash injury) showed the asymmetric distribution of studies under the funnel plot providing visual evidence of publication bias (Figure 4).

**Figure 4:**
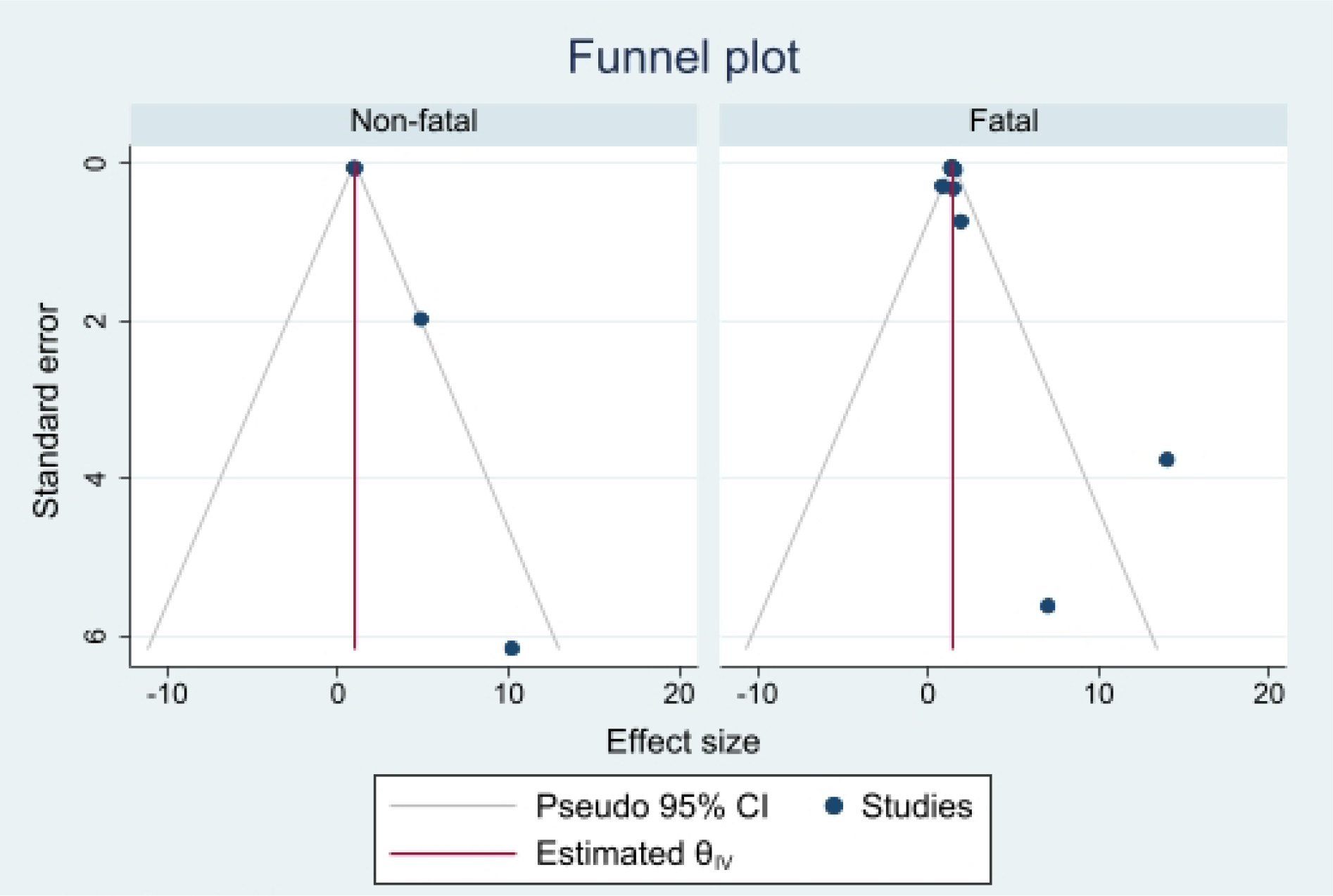
Funnel plot showing the distribution of studies in the subgroup analysis grouped by fatal and non-fatal crash injury

### 3.9 Sub-group Meta-Analysis: Morning and Afternoon Rush Hour

Four studies reported crash outcomes during the morning and afternoon rush hour periods. A total of 10 point estimates emerged from these four studies (Figure 5A). The rush-hour period was associated with a 32% increased risk of fatal and non-fatal crash injury (Pooled Risk Ratio: 1.32; 95% CI: 1.18 – 1.47). The test for heterogeneity was considerably high across all selected studies (I^2^ = 93.41%). Although most of the studies were symmetrically located around the pooled estimates, most of the studies were located outside of the margin of the funnel plot (Figure 5B). Since the margin of the funnel plot suggests where 95% of the studies would lie if there were no publication bias, the presence of some points outside of the margin made the visual interpretation of publication bias equivocal. The Egger test was not statistically significant (p=0.320), which suggests that there is no statistical evidence of publication bias. Using the Trim and Fill analysis, two studies were imputed (n=12) (Figure 5C). The adjusted effect size was 1.29 (95% CI: 1.14 – 1.43), suggesting that the rush hour period was associated with 29% increased fatal and non-fatal crash injury (Table 8).

**Figure 5:**
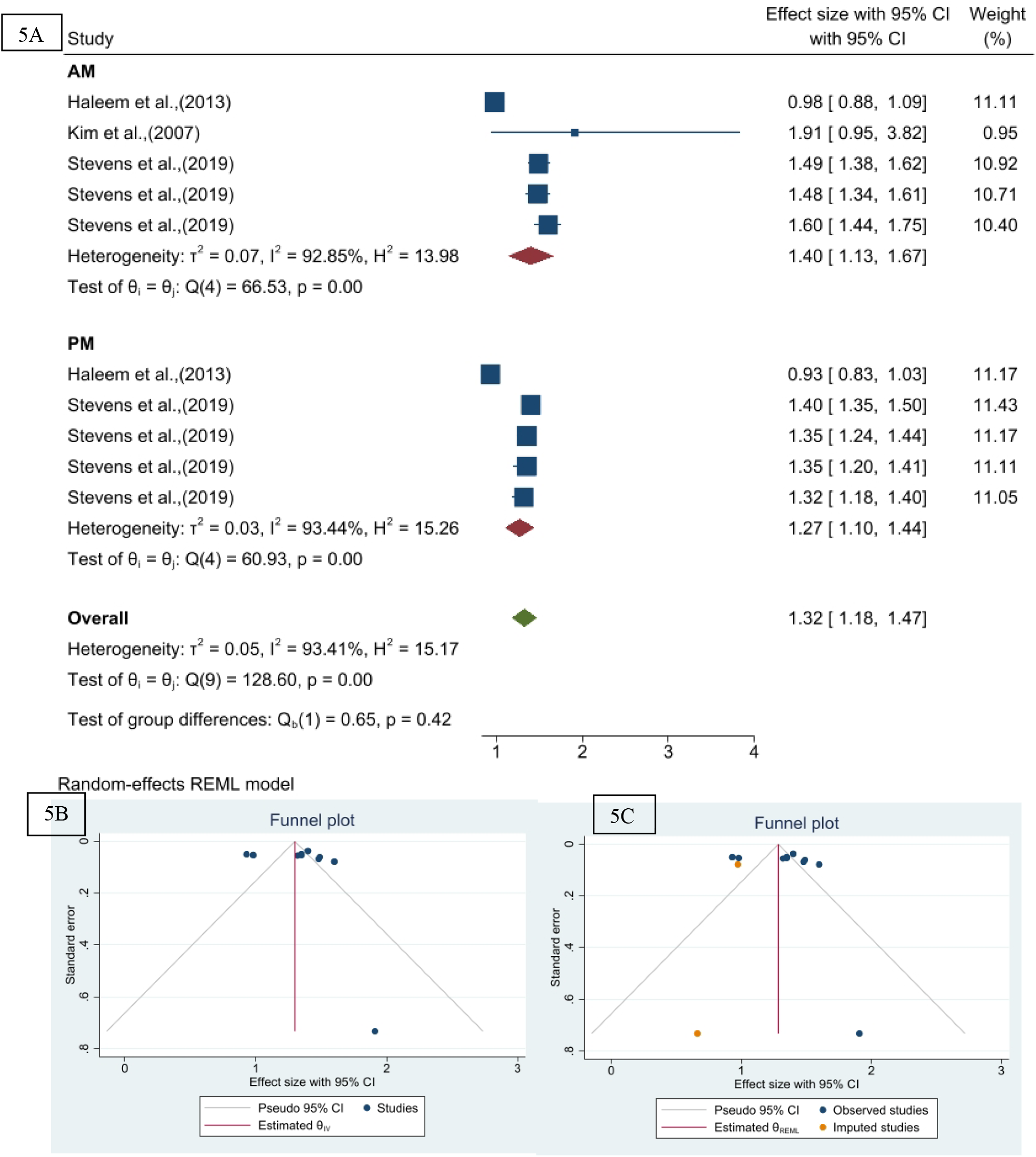
Forest plot (A) showing the sub-group analysis by morning and afternoon rush hour periods, funnel plot (B) visualizing publication bias, and Trim and Fill analysis plot (C) showing the adjustment for publication bias

**Table 8:** Summary of the Trim and Fill analysis of the sub-group analysis morning and afternoon rush hour period, showing the adjusted effect size and the Egger test evaluation of publication bias across the selected studies.

| Trim and Fill Analysis |  |  |
| --- | --- | --- |
| Studies | Theta | 95% CI |
| Observed (n=10) | 1.32 | 1.18 – 1.47 |
| Observed + Imputed (12) | 1.29 | 1.14 – 1.43 |

| Regression-based Egger test for small-study effects |  |  |
| --- | --- | --- |
| Beta (Standard Error) | Z-value | p-value |
| 1.11 (1.12) | 0.99 | 0.320 |

The subgroup analysis, measured by morning and afternoon rush hour status showed that the morning rush hour period was associated with a 40% increased risk of fatal and non-fatal crash injury (Pooled Risk Ratio: 1.40; 95% CI: 1.13 – 1.67). There was considerable high heterogeneity across the selected studies (I^2^ = 92.85%). The afternoon rush hour period was associated with a 27% increased risk of fatal and non-fatal crash injury (Pooled Risk Ratio: 1.27; 95% CI: 1.10 – 1.44). There was considerable high heterogeneity across the studies (I^2^ = 93.44%). The funnel plot of the subgroup analysis (morning and afternoon rush hour) showed the asymmetric distribution of studies under the funnel plot providing visual evidence of publication bias (Figure 6).

**Figure 6:**
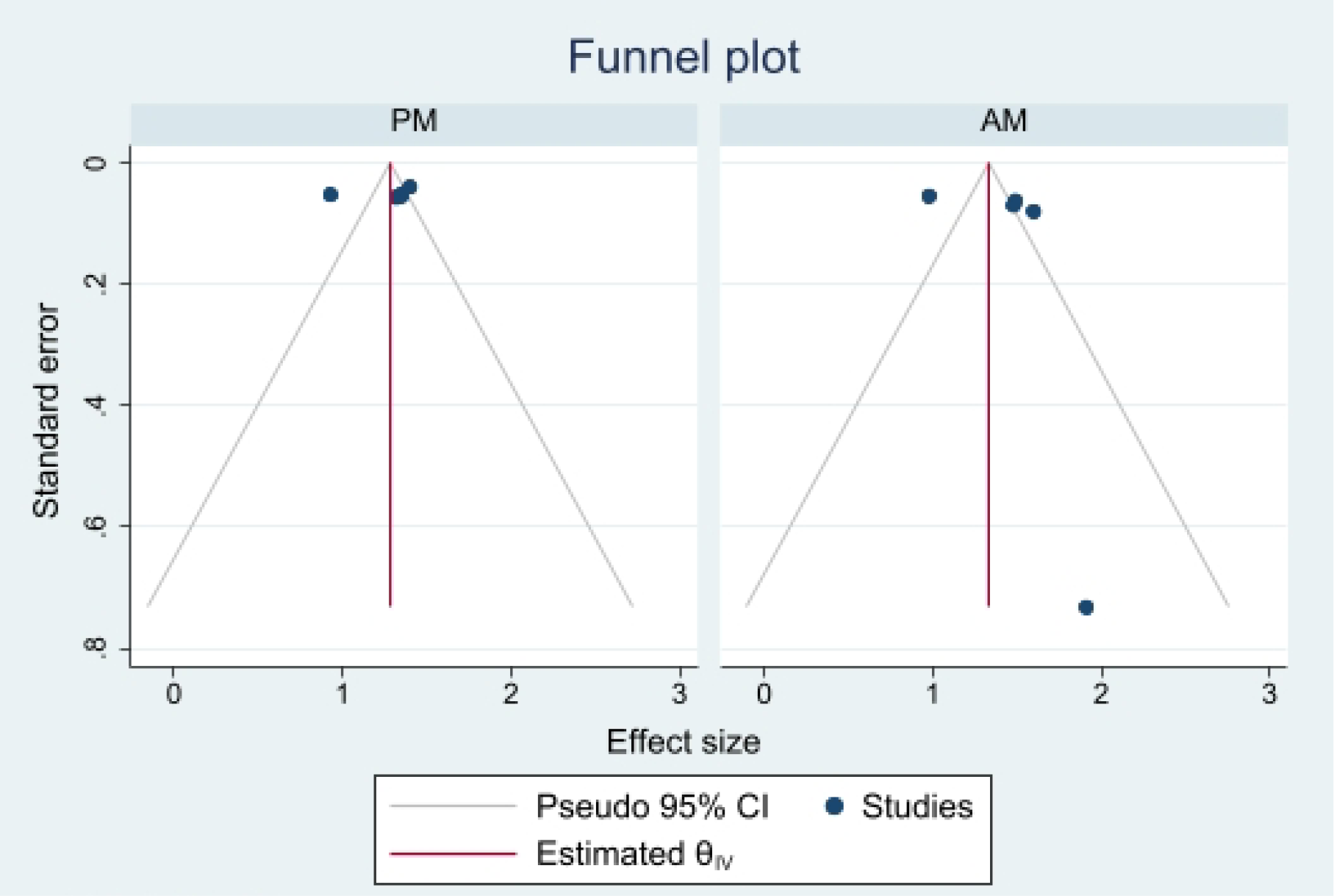
Funnel plot showing the distribution of studies in the subgroup analysis grouped by fatal and non-fatal crash injury

### 3.10 Meta-Regression Analysis

A meta-regression analysis assessed the relative risk of fatal and non-fatal crash injury by the morning and afternoon rush hour periods (Table 9). Compared to the afternoon rush hour period, the morning rush hour period was significantly associated with a 13% increased risk of fatal and non-fatal crash events (Pooled Risk Ratio: 1.13; 95% CI: 1.05 – 1.22).

**Table 9:**
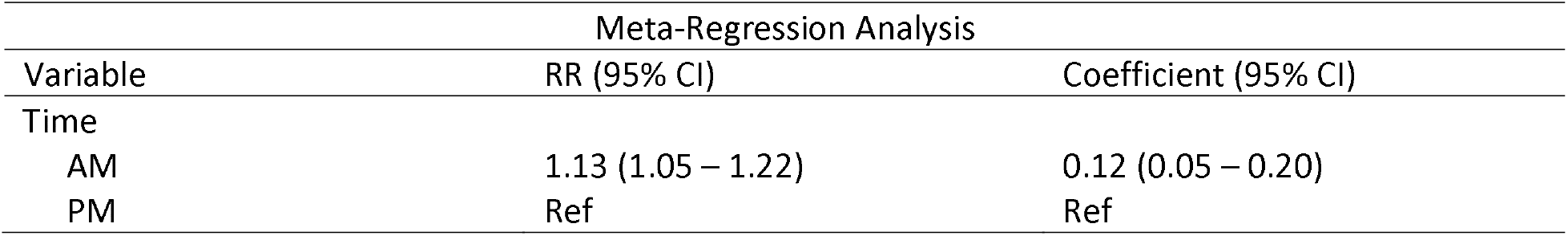
Summary of the meta-regression analysis using the rush hour categorization as the predictor.

## 4. Discussion

Across the U.S., this study estimated that the median rush hour period was from 6 to 9 am and 3 to 7 pm but the exact duration varies across states, population, population density, and county characteristics. A substantial proportion of crash events occurred during the rush hour period and rush hour traffic volume was associated with crash occurrences. Individual determinants (e.g., aggressive driving), vehicle determinants (e.g., truck driving, bicycle riding), and environmental determinants (e.g., precipitation) were associated with increased crash events or crash injuries. This study reports that the rush hour period was associated with an increased risk of fatal crash injury. The morning rush hour period was associated with increased crash injury risk compared to the afternoon rush hour period.

The rush-hour period signifies the period of “peak traffic flow” and in the U.S., two peak periods exist – the morning and afternoon rush-hour periods. Peak traffic densities vary across states, rural-urban status, county population and landmass, traffic zone regions, and time of the year. While this study identified wide-ranging rush hour periods, county-level duration in non-metropolitan and rural counties may be shorter. An earlier report from the Federal Highway Administration estimated that peak traffic flow in cities with 500,000 to one million population lasts about six hours while cities with more than one million population may have peak traffic flow lasting approximately 7.5 hours (Federal Highway Administration, 2017).

While defining the rush hour period using discrete-time points has its advantages, the rush hour period is not static. Defining rush hour as a predictor of crash events may require quantifying the volume of traffic flow during the rush hour period, assessing the presence (or absence) of the variable of interest in the rush hour period, or using the rush hour period as an effect modifier, stratification, or interaction variable, if statistical criteria are met. Introducing indices of the rush hour period into analytical models may inform decisions on the appropriateness of using the rush hour period as a proxy for intervention. Additionally, understanding how the rush hour period associates with predictors of crash events or crash injury may inform policy on driving behavior, road environmental designs, and designs of safer vehicles, including self-driving cars.

While there exist significant fatal crash injuries during the morning and afternoon rush hour periods, the causes of such crash injuries may differ. The morning rush hour period represents the time most road users are rushing to their workplace. The afternoon rush hour period mirrors the period road users leave work to their homes or secondary destinations. Earlier studies have reported increased texting during the afternoon rush hour period compared to other times of the day (Drivemode Research Lab, 2018). While speeding may or may not be associated with the rush hour period depending on the type of road and the volume of vehicles, drivers are more likely to speed during the morning rush hour period than at any other time of the day (Richard et al., 2012). This driving behavior may explain the increased proportion of weather-related crashes during the morning rush hour period compared to the afternoon rush hour period (Call et al., 2019).

A few studies reported non-significant relationships between rush hour traffic volume and crash event/injury (Das et al., 2008; Haleem & Gan, 2013; Lee et al., 2007). Conceptually, during the rush hour period, there should be reduced speed due to the increased density of vehicles on the road. However, this assumption may hold if the road type is either arterial roads or freeways. An earlier report by the Federal Highway Administration reported that crash fatality rates, measured at all times of the day, were highest on collectors and local streets and least on interstate and arterials (Federal Highway Administration, 2000). Studies selected in this review that assessed crashes on arterial roads and freeways produced either reduced risks of crash events/injuries or non-statistical associations (Das et al., 2008; Haleem & Gan, 2013; Lee et al., 2007). This may suggest that rush hour-related crash injuries may be less of a problem on roads with heavy traffic such as arterial roads and freeways and may be related to local streets and collectors.

Rush hour-related crashes are an under-researched area in traffic injury prevention. Thirteen papers assessed the relationship of rush hour, measured in any form, and crash injury or event. Eight of the 13 papers assessed the rush hour as a control variable, while the other studies used the rush hour period as a stratification or interaction variable or for descriptive purposes. Seven of the 13 papers were published within the last decade. None of the studies assessed the rush hour period or rush hour traffic volume as the main predictor of crash event/injury. While a few studies assessed the individual, vehicle, and environmental determinants of crash injury during the rush hour, research in the domains aimed at achieving zero fatality (Ecola et al., 2018; Federal Highway Administration, 2020) such as speeding, distracted driving, seatbelt use, road environmental infrastructure, and crash response, are missing. Additionally, no study evaluated the role of the social determinants of health (Healthy People, 2020) in the occurrence of fatal and non-fatal crash injuries in the U.S. Further, none of the studies used spatial or spatiotemporal estimators – tools that may adjust for spatial and temporal autocorrelation and provide information of cluster location.

Crash event and crash injury represent two different concepts that are aimed at achieving a single goal – injury prevention. A crash event refers to the presence of solitary or multiple collisions with or without the presence of injury or loss of lives. Crash injury captures the occurrence of the collision and the presence or absence of fatal or non-fatal injury. Also, there exists a hierarchical difference in the crash event and crash injury conceptualization. A crash event (count or occurrence) may be conceptualized at the community level while crash injury may be measured at the individual level. Aggregating either measures of crash outcome to provide county, zip code or state-level estimates may require defining appropriate standardization which may be population estimates (Marquis & Wang, 2015) or traffic volume (Abdel-Aty, Pemmanaboina, & Hsia, 2006; Lee et al., 2007; Mitra & Washington, 2012). While the NHTSA reports counts and rates of fatal and non-fatal crash injuries (National Center for Statistics and Analysis, 2019b; National Highway Traffic Safety Administration, 2018, 2019), information on weighted or adjusted estimates of fatal and non-fatal crash injuries would provide additional information on how the crash rates vary by states, across years, and by specific predictors.

This study has several limitations. All the studies employed a retrospective study design, and the observed relationship of the rush hour and crash injury does not infer causation. This study reports statistical evidence of publication bias, which may be due to earlier studies attenuating the report of non-significant variables (Das et al., 2008; Duddu et al., 2019). Also, there is a possibility that some publications were systematically missed as these articles might not have been indexed in the databases that were searched. The inherent weakness of the primary studies is reflected in this meta-analysis as issues relating to analytical bias, coverage and sampling errors, and misclassification of the rush hour period in the original studies might influence this study’s results. Despite these limitations, this study represents one of the few studies that highlight the crash injury risks associated with the rush hour period. Additionally, this study identifies the rush hour period as an area needing more scholarly research and a potential proxy for different crash injury interventions.

## 5. Conclusion

In summary, a substantial proportion of crash injuries occur during the rush hour period in the U.S. The morning and afternoon rush hour periods are each associated with an increased risk of fatal crash injuries in the U.S. As the U.S journeys towards achieving zero fatal deaths, there is a need for more scholarly research on the rush hour-related crash injury pattern. The knowledge of the pattern of crash injuries, as it varies across states, rurality/urbanicity, and counties may inform policy and the design of interventions. This systematic review and meta-analysis further demonstrate the paucity of information on the individual, environmental, and community-level crash injury determinants during the rush hour period. Increased knowledge of the pattern of rush hour-related fatal and non-fatal crash injuries may inform policies related to rush hour driving, resource allocation, and targeted interventions.

## Data Availability

This study is a systematic review and meta-analysis. The data used are from the studies selected in the study.

## Acknowledgment

The authors acknowledge Tasha Gills, MPH, and Jessica Hoyle, MMT for their editorial assistance.

